# Bridging the polar and hydrophobic metabolome in single-run untargeted liquid chromatography-mass spectrometry dried blood spot metabolomics for clinical purposes

**DOI:** 10.1101/2021.03.22.21254119

**Authors:** Hanne Bendiksen Skogvold, Elise Mørk Sandås, Anja Østeby, Camilla Løkken, Helge Rootwelt, Per Ola Rønning, Steven Ray Wilson, Katja Benedikte Prestø Elgstøen

**Affiliations:** National Unit for Screening and Diagnosis of Congenital Pediatric Metabolic Disorders, Department of Medical Biochemistry, Oslo University Hospital, Sognsvannsveien 20, 0372 Oslo, Norway; Department of Mechanical, Electronic and Chemical Engineering, Faculty of Technology, Art and Design, Oslo Metropolitan University, Pilestredet 35, 0166 Oslo, Norway; Department of Chemistry, University of Oslo, Sem Sælands vei 26, 0371 Oslo, Norway; Hybrid Technology Hub-Centre of Excellence, Institute of Basic Medical Sciences, Faculty of Medicine, University of Oslo, Domus Medica, Gaustad, Sognsvannsveien 9, 0372 Oslo, Norway

**Author notes:** Corresponding author: Katja Benedikte Prestø Elgstøen.

**Keywords:** Metabolomics, Dried blood spots, LC-MS, Inborn errors of metabolism

## Abstract

Dried blood spot (DBS) metabolite analysis is a central tool for the clinic, e.g. newborn screening. Instead of applying multiple analytical methods, a single liquid chromatography-mass spectrometry (LC-MS) method was developed for metabolites spanning from highly polar glucose to hydrophobic long-chain acylcarnitines. For liquid chromatography, a diphenyl column and a multi-linear solvent gradient operated at elevated flow rates allowed for an even-spread resolution of diverse metabolites. Injecting moderate volumes of DBS organic extracts directly, in contrast to evaporation and reconstitution, provided substantial increases in analyte recovery. Q Exactive MS settings were also tailored for sensitivity increases, and the method allowed for analyte retention time and peak area repeatabilities of 0.1-0.4 % and 2-10 %, respectively, for a wide polarity range of metabolites (logP −4.4 to 8.8). The method’s performance was suited for both untargeted analysis as well as targeted approaches, evaluated in clinically relevant experiments.

**Graphical Abstract:** 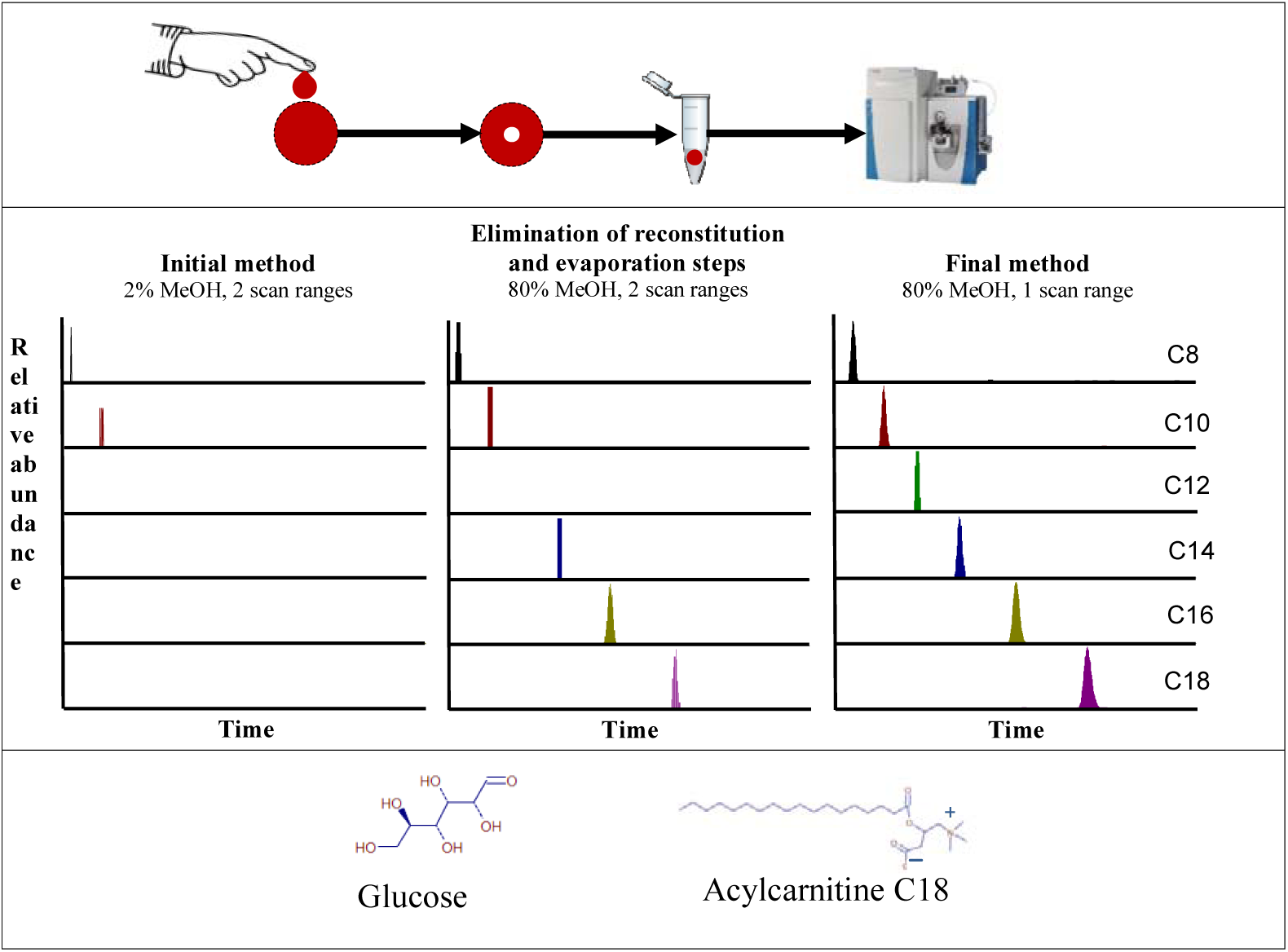

## 1 Introduction

Reliable and accurate analyses of biological samples are essential for clinical purposes. Untargeted analyses of small biomolecules, i.e. metabolomics, provide the advantage of enabling the detection of large numbers of compounds at the same time. The approach has been described as having no discrimination in terms of which compounds that can be detected within certain instrumental limitations, e.g. mass range [1,2]. In human clinical samples, these metabolites include endogenous molecules, all the exogenous molecules we are voluntarily and involuntarily exposed to, as well as a myriad of metabolites generated by our microbiomes.

Inborn errors of metabolism (IEMs) represent a large and diverse group of diseases [3-5]. An IEM is typically caused by genetic mutations in a single gene, leading to changes in function or quantity of a vital enzyme. This in turn leads to deviations in the patient’s metabolism. IEMs can have serious consequences such as severe brain damage or death, and are included in newborn screening programs in most countries. For IEM diagnostics, targeted analyses are mostly used, i.e. monitoring a limited number of predefined analytes. Untargeted analyses can also be of great importance for these diseases, as symptoms are often diverse and unspecific, making diagnostics based on targeted analyses difficult and potentially very time consuming [5].

Dried blood spots (DBS) are increasingly used in diagnostics due to ease of sampling, storage, and transportation, as well as higher stability of most analytes compared with e.g. plasma or whole blood samples [6-8]. Palmer et al. reported detecting more metabolites in DBS than in plasma samples, likely due to red and white blood cell metabolites [9]. DBS are also widely used in newborn screening [10,11], allowing newborn screening even in remote areas, as samples can be gathered anywhere in the world and sent to laboratories elsewhere for analysis.

Mass spectrometry (MS) is a key tool in metabolomics [12-14]. MS enables reliable and accurate identification of compounds, and generally provides increased sensitivity compared to e.g. NMR spectroscopy techniques [15]. Regarding clinical metabolic screening, methods may be built upon direct infusion mass spectrometry due to e.g. speed and simplicity [10,16]. On the other hand, applying liquid chromatography (LC) upstream to MS has the advantage of providing additional separation of compounds, thereby increasing the number of identifiable compounds and reduction of matrix effects such as ion suppression and ion enhancement [14,17]. Applying LC-MS also increases the chance of identification of isomers and provides the additional parameter of retention time for identification purposes [14,18].

A key challenge with analyses of biological samples is the wide polarity range of clinically relevant metabolites, ranging from e.g. hydrophobic fatty acids to polar amino acids, often addressed by using several analytical methods/platforms e.g. employing gas chromatography in addition to hydrophilic interaction liquid chromatography (HILIC) and reversed phase LC, to cover a broad part of the metabolome [19-23]. Although comprehensive analysis is provided, multi-separation approaches can be time consuming and laborious, sometimes requiring extensive sample preparation procedures. Several single metabolomics methods have been developed for e.g. plasma and urine analysis, with excellent results, see e.g. references [24,25]. Broad range single methods are even beginning to merge different omics; He et al. recently combined lipidomics and proteomics with a single-shot technology [26]. We set out to develop and optimize a single method suited for covering a broad range of metabolites focusing on dried blood spots, while still ensuring simplicity for practical clinical use, e.g. IEM diagnostics. We were interested in if a single LC-MS method would have satisfactory analytical performance (for example, robust retention times and peak area measurements) for metabolites ranging from sugars to lipids.

In this work, we present optimizations and demonstrations of a metabolomics LC-Orbitrap MS method for DBS analysis intended to “bridge” the hydrophobic and polar metabolome in a single run. We focus on factors including retention time/peak area stability, peak capacity, DBS extraction and MS parameter settings. We demonstrate the method for both untargeted analysis and a targeted approach.

## 2. Experimental

### 2.1 Equipment

Filter paper cards used were Whatman 903 Protein Saver cards (GE Healthcare Life Sciences, Chicago, IL, USA). A manual puncher from McGill (Advantus corp., Jacksonville, FL, USA) was used to punch the DBS. Micro tubes were obtained from Sarstedt (Nümbrecht, Germany). For extraction, a Thermomixer Comfort (Eppendorf, Hamburg, Germany) was used. Glass tubes for evaporation to dryness were obtained from VWR (Radnor, PA, USA), and the evaporator used was a TurboVap LV (Caliper Life Sciences, Waltham, MA, USA). HPLC vials, caps and inserts were from La-Pha-pack (Thermo Scientific, Waltham, MA, USA).

The following LC columns were evaluated: Polaris C18-Ether and Pursuit XRs Diphenyl (both from Agilent Technologies (Santa Clara, CA, USA)), C18-Pentafluorophenyl (PFP) from ACE Technologies (Aberdeen, Scotland), Aeris Peptide XB-C18 (Phenomenex (Torrance, CA, USA)), and Raptor Biphenyl (Restek (Bellefonte, PA, USA)). For column specifications, see **Table 1**.

**Table 1.** Specifications of evaluated columns

| Column | Length, mm | Diameter, mm | Particle size, $\mu\text{m}$ | Pore size, $\text{\AA}$ | Surface area, $\text{m}^2/\text{g}$ | Carbon load, % |
| --- | --- | --- | --- | --- | --- | --- |
| Polaris C18-Ether | 250 | 2.0 | 3.0 | 180 | 200 | 12 |
| Pursuit XRs Diphenyl | 250 | 2.0 | 3.0 | 100 | 440 | 15 |
| ACE C18-PFP | 250 | 2.1 | 3.0 | 100 | 300 | 14 |
| Aeris Peptide XB-C18 | 250 | 2.1 | 3.6 | 100 | 200 | 10 |
| Raptor Biphenyl | 150 | 2.1 | 2.7 | 90 | 150 | 7 |

### 2.2 Chemicals and solvents

All water used was of type 1 (>18 MΩ cm), obtained from MilliQ ultrapure water purification system (Merck Millipore, Darmstadt, Germany). Methanol was obtained from Rathburn Chemicals (Walkerburn, Scotland). Formic acid (98%) was obtained from Merck.

Tobramycin, acylcarnitines C2, C12, and C16, D2 glycolic acid, D6 glucose and acylcarnitines D3 C2, D3 C12 and D3 C16 were obtained from Larodan (Solna, Sweden). D4 succinic acid, ^13^C creatine and uric acid were purchased from Sigma (Darmstadt, Germany). Vancomycin (1000 mg powder) was obtained from MIP Pharma GmbH (Blieskastel, Germany). ^13^C_2_ guanidinoacetate was obtained from Dr. H Ten Brink (VU University Medical Center, Amsterdam, The Netherlands). Creatinine was obtained from Merck. Creatine was obtained from Nutritional Biochemical Corporation (Cleveland, OH, USA).

### 2.3 Method development

The following parameters were evaluated and optimized. MS parameters, with regards to signal intensity: electrospray voltage (evaluated values: 1, 2, 3.5, 4, 5, 6, and 7 kV). Electrospray needle position (evaluated positions: A-D, A being the closest to the inlet). Resolution (evaluated values: 17 500, 35 000, 70 000, and 140 000 FWHM (at *m/z* 200)). Automatic gain control (AGC) target value (evaluated values: 2E4, 5E4, 1E5, 2E5, 5E5, 1E6, 3E6, and 5E6 ion counts). Evaluation of using either a broad (and split) scan range of *m/*z 50-750 and 750-1700 or only one range of *m/*z 50-750 was done with regards to peak area.

LC parameters, with regards to peak distribution: LC column (see **Table 1** for names and specifications) and gradient elution profile (see **Figure 1** for evaluated profiles; more details are described below). LC parameters, with regards to peak capacity: injection volume (evaluated volumes: 2, 10 and 20 µL) and mobile phase flow rate (evaluated rates: 150 and 300 µL/min).

**Figure 1.**
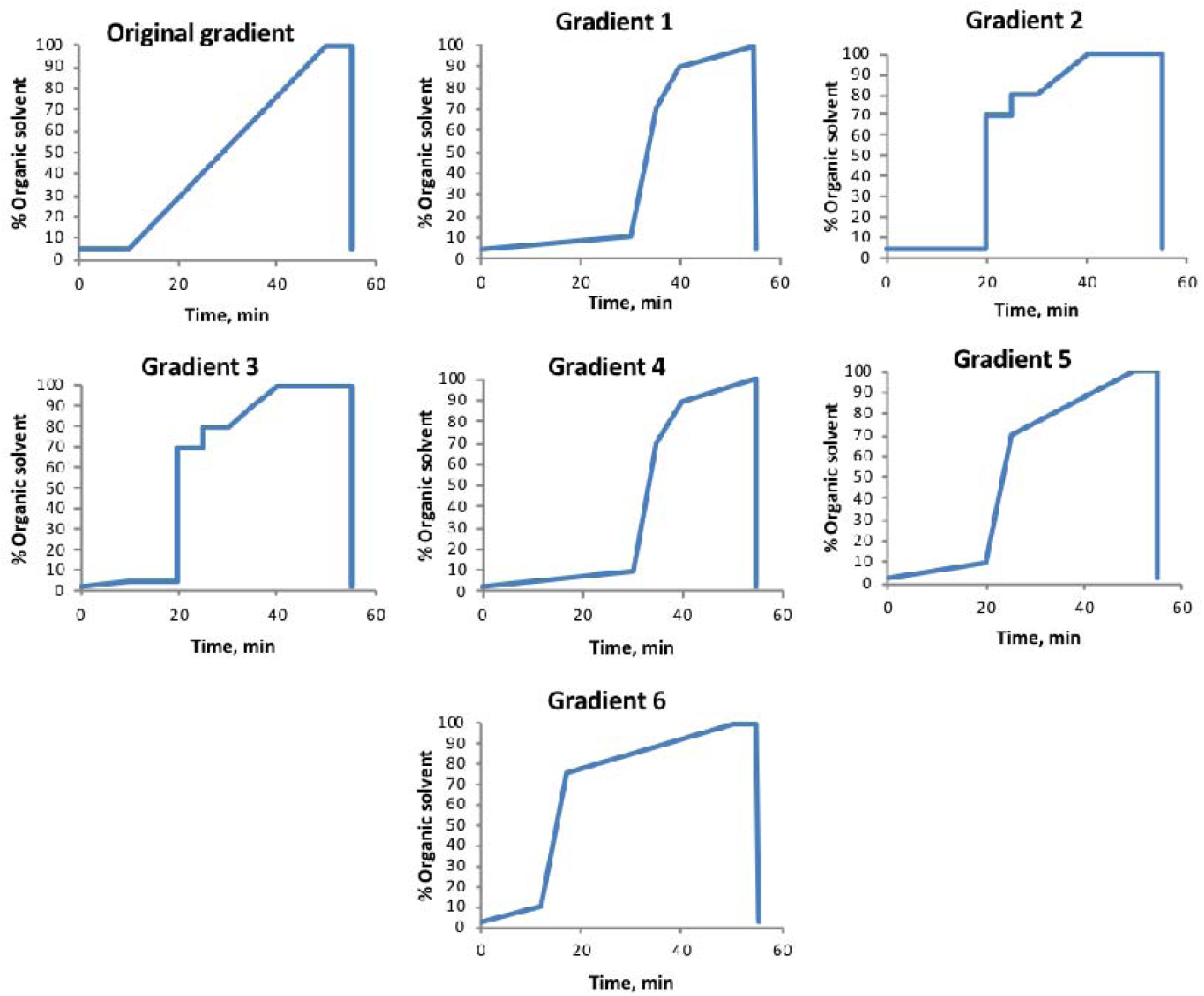
Gradient profiles tested. Mobile phase A: water with 0.1% formic acid. Mobile phase B: methanol with 0.1% formic acid.

Figure 1. shows the evaluated gradient elution profiles (evaluated using Pursuit XRs Diphenyl column). The original gradient was the starting point of the gradient profile optimization, while gradients 1-6 were defined based on when compounds in a spiked DBS eluted. Total analysis time was later reduced to 32.5 minutes.

Sample preparation optimization: evaluation of evaporating samples to dryness and re-solving in 2, 40 and 80% methanol, respectively, versus injection of 80% methanol extract without evaporation to dryness, with regards to peak area. DBS punch location (evaluated locations: A-D, A being in the center of the spot, D in the perimeter (see **Figure 6** in **Results**), with regards to standard deviation of measured peak area (10 spots were used, providing 10 punches from each location).

### 2.4 Sample preparation

For optimization experiments, whole blood from a healthy volunteer was used. The whole blood was either spotted onto a filter paper card directly, or mixed with aqueous standards (50:50 v/v) and spotted onto a filter paper card. The same blood sample was used in all experiments for optimization of a chosen parameter. Whole blood was either mixed with the standards directly before spotting, or stored in a freezer at −80° C and thawed before mixing and spotting.

Dried blood spots were either made immediately prior to each experiment, or prepared samples were stored at −80°C before use. The following steps constitute the final sample preparation: 3.2 mm punches were punched from DBS (∼ 3 µL whole blood or ∼ 1.5 µL whole blood for the samples consisting of whole blood mixed with aqueous standards), and extracted in a micro tube with 100 µL 80% aqueous methanol with 0.1% formic acid using a thermomixer for 45 minutes (at 45°C, 700 rpm). Samples were transferred to an HPLC vial for analysis directly after extraction.

### 2.5 Instrumentation

LC instrumentation used was Dionex Ultimate 3000 UHPLC system pump, column department and autosampler, from Thermo Scientific. The MS used was a Q Exactive Orbitrap (Thermo Scientific). The ionization source was an electrospray, and samples were analyzed in both positive and negative mode (in separate injections).

### 2.6 Settings and details

The following settings constitute the final method. LC column: Pursuit XRs Diphenyl (see **Table 1** for details). Injection volume: 2 µL. Mobile phase (A: water with 0.1% formic acid, B: methanol with 0.1% formic acid) flow rate: 300 µL/min. Gradient elution profile: profile 6 in **Figure 1**. Column temperature was 30°C, and total analysis time was 32.5 minutes. Re-equilibration time was 10 minutes.

Scan type was Full MS (scan range *m/z* 50-750). Resolution: 70 000 FWHM (at *m/z* 200). AGC target value: 1 000 000 ion counts. Maximum injection time: 250 ms. Electrospray settings: sheath gas (N_2_) flow rate: 40 (a.u.), auxiliary gas (N_2_) flow rate: 10 (a.u.), sweep gas (N_2_) flow rate: 2 (a.u.), capillary temperature: 250°C, S-lens RF level: 50, auxiliary gas heater temperature: 300°C, electrospray voltage: 3.5 kV, electrospray needle position: C.

Equation 1 was used for calculation of peak capacity (P_c_).

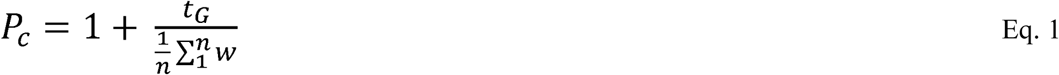

Where t_G_ is the gradient elution time, n is the number of peaks, and w is the peak width at baseline (13.4% peak height) for each peak.

### 2.7 Computer software

Software used was Xcalibur (Version 4.2.47), Tune (version 2.11), and SII for Xcalibur 1.5, all from Thermo Scientific. Compound Discoverer 2.1 (Thermo Scientific) was used for data processing.

### 2.8 Approval by Regional Committees for Medical and Health Research Ethics

The use of whole blood from healthy volunteers was approved by the Regional Committees for Medical and Health Research Ethics (case number: 173346).

## 3 Results and Discussion

An LC-Orbitrap MS method for metabolomics analyses of DBS was optimized regarding sample preparation, chromatographic properties, and MS conditions for coverage of a broad range of metabolites and a high degree of sensitivity. A selection of endogenous and isotopically labelled metabolites (spiked in controlled amounts) was used for the optimization experiments. Included in this list were hydrophobic acylcarnitines (key biomarkers in newborn screening) as well as more polar amino acids including valine and tyrosine (biomarkers of maple syrup urine disease and tyrosinemia, respectively). Using the same methods, proof-of-concept demonstrations were performed for both targeted and untargeted DBS applications. Below is a more detailed presentation of the optimizations, method evaluation, and proof-of-concept experiments.

### 3.1 Method development

#### 3.1.1 MS optimization

Q Exactive mass spectrometry parameters were optimized for metabolites from DBS with regards to signal intensity. For optimization of MS parameters, we used a standard mix of metabolites (about 5 µmol/L each) with a range of molecular weight, structure and polarity, see **Figure 2**. In these experiments, all monitored standards were isotopically labelled to ensure that observed changes in signal intensities were caused only by parameter settings without interference from endogenous contributions (except for the drug vancomycin). Vancomycin was added to include a compound with a relatively large mass that is also analyzed in our routine laboratory, making method comparison possible. Optimization experiments were primarily performed using an aqueous mix of standards mixed with whole blood (50:50 v/v) and spotted onto a filter paper card. For these samples, the following MS settings were considered to be the best choices: electrospray voltage 3.5 kV, electrospray needle position C (options were A-D, A being the closest to the inlet, with a difference between the positions of approximately 3.5 mm), resolution 70 000 FWHM (at *m/z* 200), and automatic gain control (AGC) target value 1 000 000 ion counts. See **Figure 2** for comparison to other settings. AGC target values and electrospray voltage were of highest significance regarding sensitivity. Importantly, sensitivity increased significantly (92-109% increase in peak area of investigated compounds) when changing from a split scan range (*m/z* 50-750, 750-1700) to one scan range (*m/z* 50-750).

**Figure 2.**
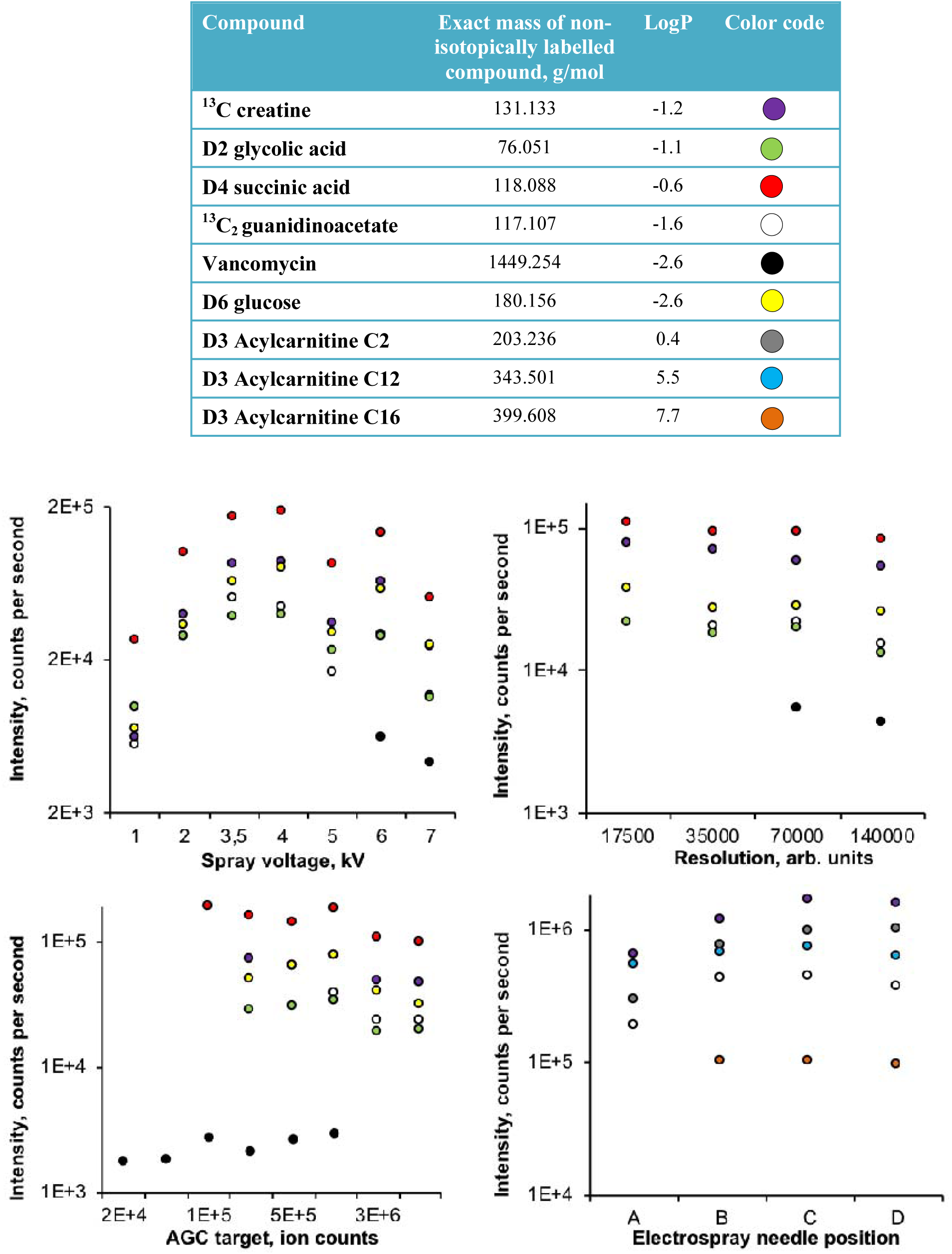
**Top:** Exact mass and logP of compounds used for MS optimization experiments. Exact mass was calculated by the Tune software from Thermo Scientific. LogP values were found at Pubchem [27]. Colored dots show the color of each compound in the plots. **Bottom:** Peak intensity with tested settings.

#### 3.1.2 LC optimization

Chromatographic parameters were optimized for metabolites from DBS firstly with regards to peak distribution and peak capacity. In these experiments, the monitored standards were non-isotopically labelled metabolites but related to those in the MS experiments, see **Figure 3**. Optimization experiments were primarily performed using an aqueous mix of standards mixed with whole blood (50:50 v/v) and spotted onto a filter paper card. The following LC parameter settings were considered to be the best choices (see **Figure 3** for comparison to other settings): analytical column: Pursuit XRs Diphenyl (hydrophobic + pi-pi interactions, provided increased separation of polar analytes), multi-linear gradient elution profile 6 in **Figure 1** (interpreted as providing the most even elution distribution of investigated compounds), injection volume 2 µL (associated with the highest peak capacity, e.g. due to not overloading the column), and mobile phase flow rate 300 µL/min (no large difference in peak capacity between tested flow rates, allowing a reduction of the analysis time)

**Figure 3.**
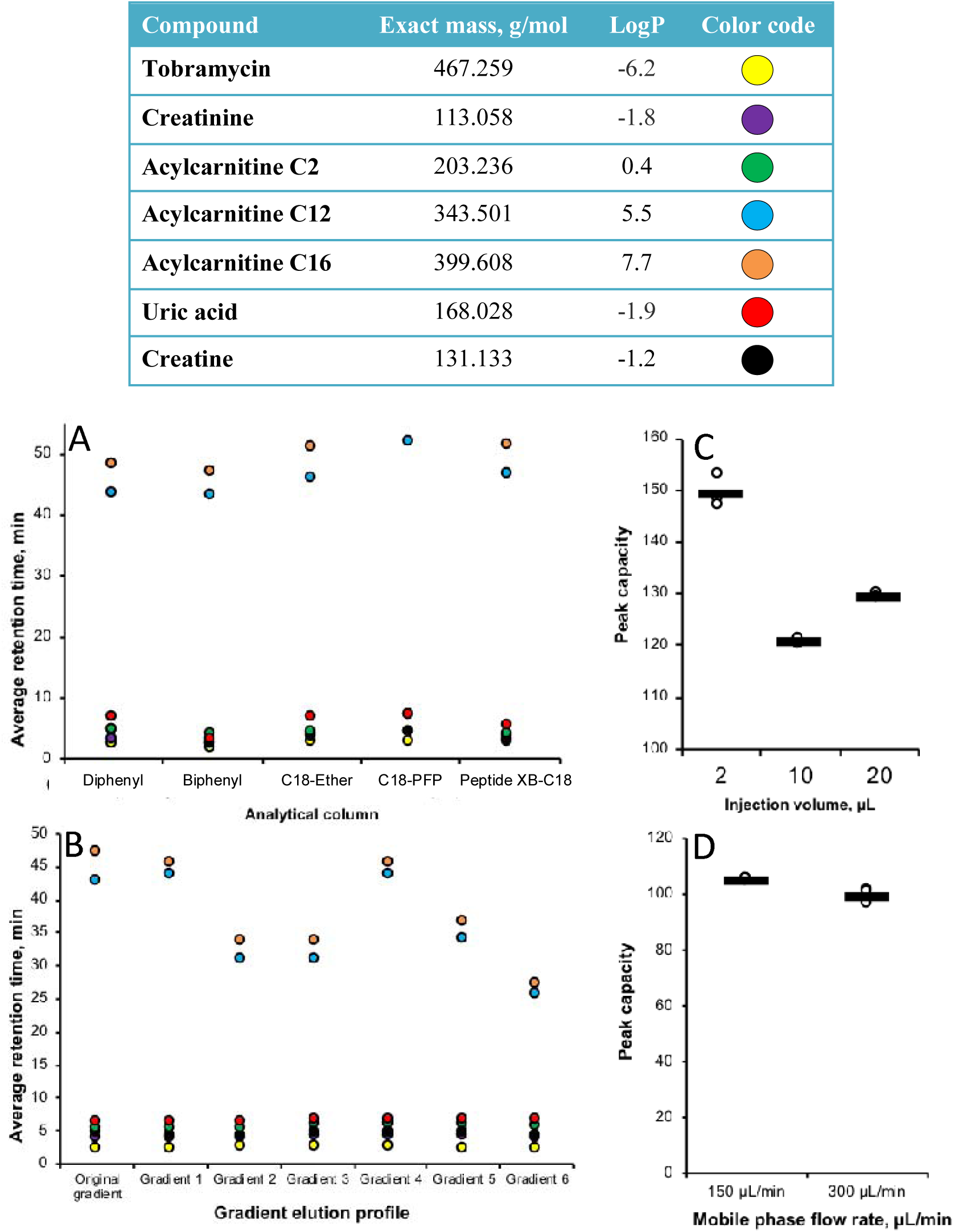
**Top:** Exact mass and logP of compounds used for LC optimization experiments. Exact mass was calculated by the Tune software from Thermo Scientific. LogP values were found at Pubchem [27]. Colored dots show the color of each compound in the plots. **Bottom:** average retention time (n=3) of compounds for tested columns (A) and tested gradient elution profiles (B), and average peak capacity (n=3) for tested injection volumes (C) and tested mobile phase flow rates (D).

#### 3.1.3 Sample preparation optimization

Sample preparation parameters were optimized for metabolites from DBS with regards to recovery/detection capability. The following steps and settings constitute the final sample preparation protocol: extraction of one punch from a DBS with 100 µL 80% aqueous methanol with 0.1% formic acid, and thermo-mixing for 45 minutes at 45°C (700 rpm). The solution was directly transferred to an HPLC vial, as this gave improved recovery and detection (28-40% increase) for all compounds tested compared to evaporating to dryness and re-solving in the starting mobile phase (**Figure 4)**. The simplified procedure of injecting higher organic contents did not lead to substantial changes in retention time for any of the compounds investigated.

**Figure 4.**
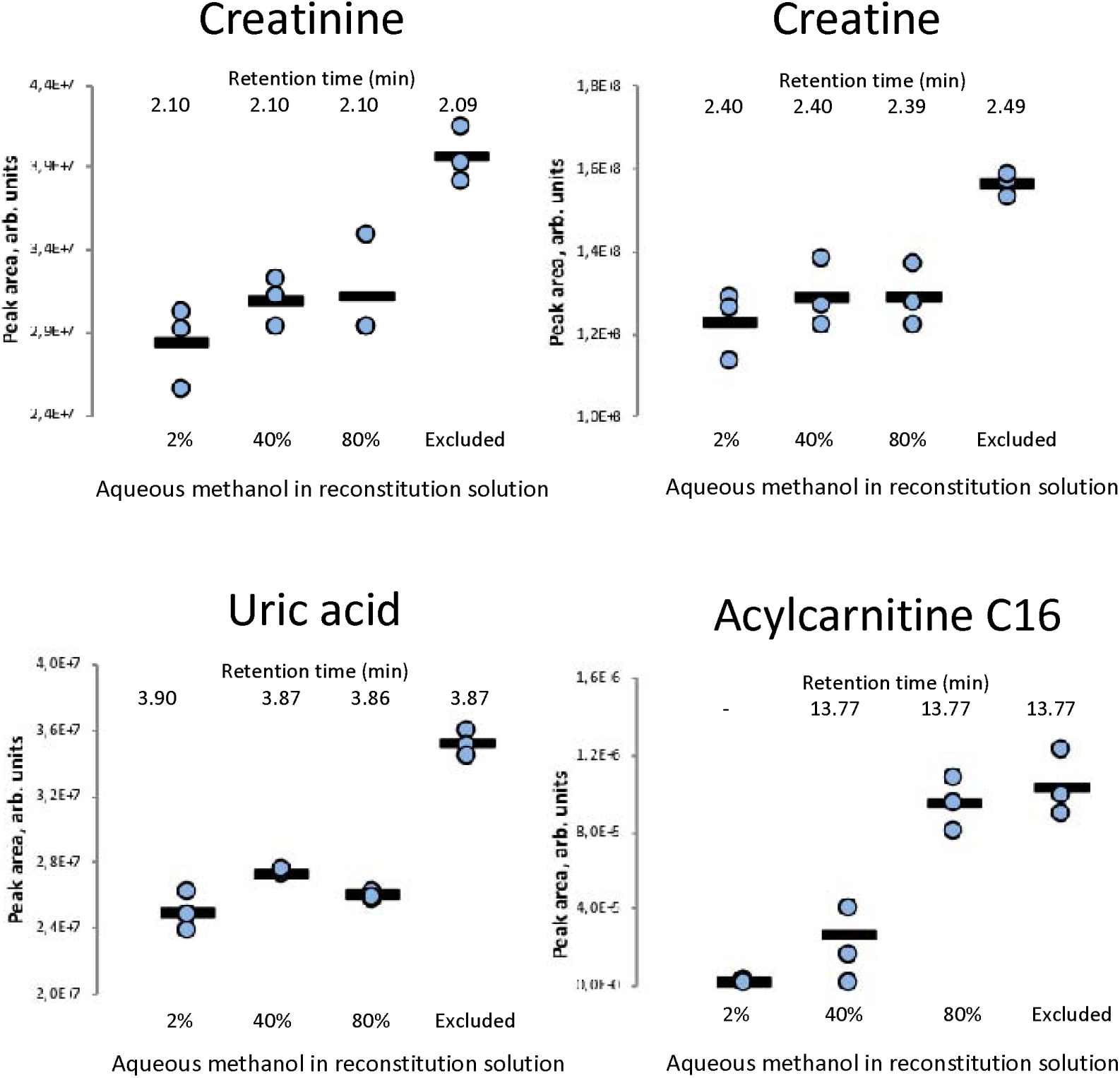
Improved sensitivity with increased amount of methanol. Peak area in DBS samples prepared with 2%, 40% and 80% aqueous methanol in the reconstitution solution, and with the exclusion of the reconstitution and evaporation steps. Retention times of the compounds for each organic solvent concentration are shown.

As shown in **Figure 5**, eliminating the evaporation to dryness step and changing from a broad and split (*m/z* 50-750, 750-1700) to a narrow (*m/z* 50-750) scan range significantly improved detection of acylcarnitines.

**Figure 5.**
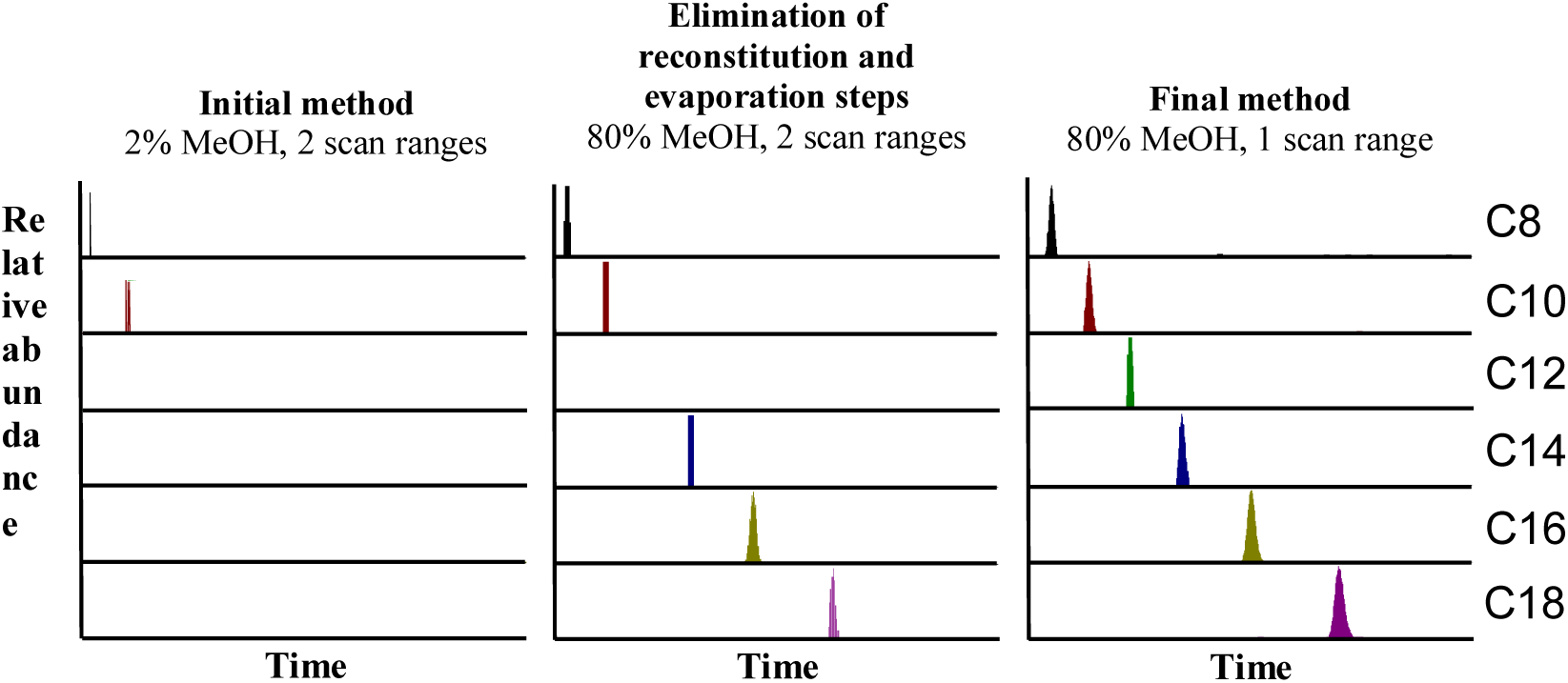
Extracted ion chromatograms of a selection of acylcarnitines showing detection improvement when changing from evaporation to dryness and a broad and split scan range (*m/z* 50-750, 750-1700) to direct transfer of extract to HPLC vial and a narrow scan range (*m/z* 50-750).

To evaluate potential differences in punch location within the DBS, four punch locations were investigated (see **Figure 6**). We observed a larger relative standard deviation (RSD %) in punches taken from the perimeter of the spot compared to the center. Center punches were thus considered to be the best choice.

**Figure 6.**
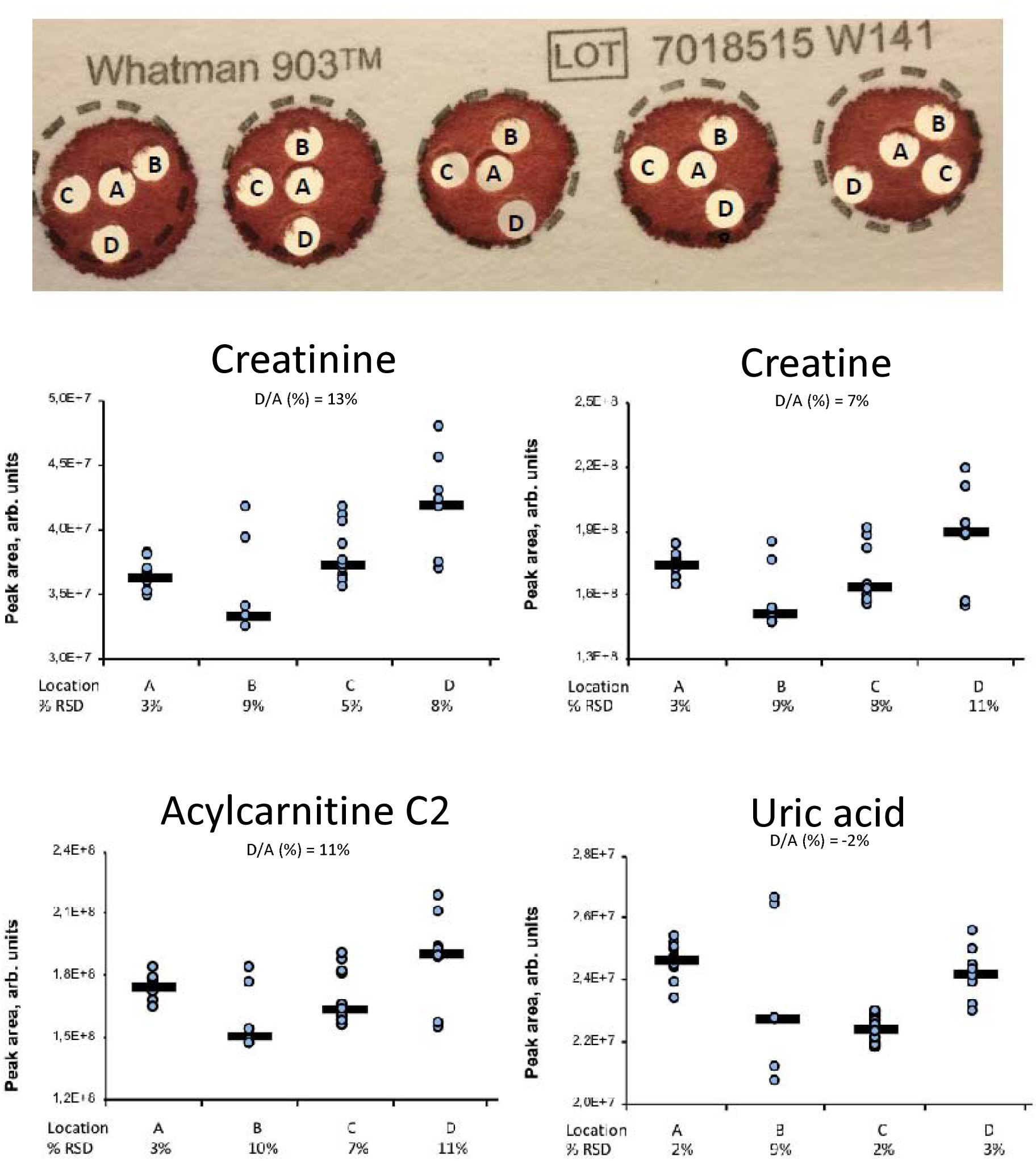
**Top:** Punch locations tested. 10 spots were used, providing 10 punches from each location. **Bottom:** Peak areas with RSD % of selected compounds in DBS samples prepared with punches taken from locations A, B, C and D.

### 3.2 Evaluation of peak area linearity

To evaluate the abilities of our DBS MS platform, we investigated the effect on measured peak area of endogenous metabolites when increasing the number of DBS punches (1, 2, 3 and 4 punches, equivalent to about 3, 6, 9 and 12 µL whole blood, respectively). Peak area linearity was overall satisfactory for the investigated analytes. As shown in **Figure 7** with glycine and alanine as examples, peak area increased linearly with increasing number of punches (R^2^ ranging from 0.9358 = ornithine, to 0.9994 = alanine). To monitor instrument performance and repeatability, the same DBS sample (healthy volunteer) was injected three times each day during an analysis run of 11 days. **Table 1** (**supplementary material**) shows a high repeatability regarding average retention time and peak area (0.1-0.4%, 2-10%, respectively).

**Figure 7.**
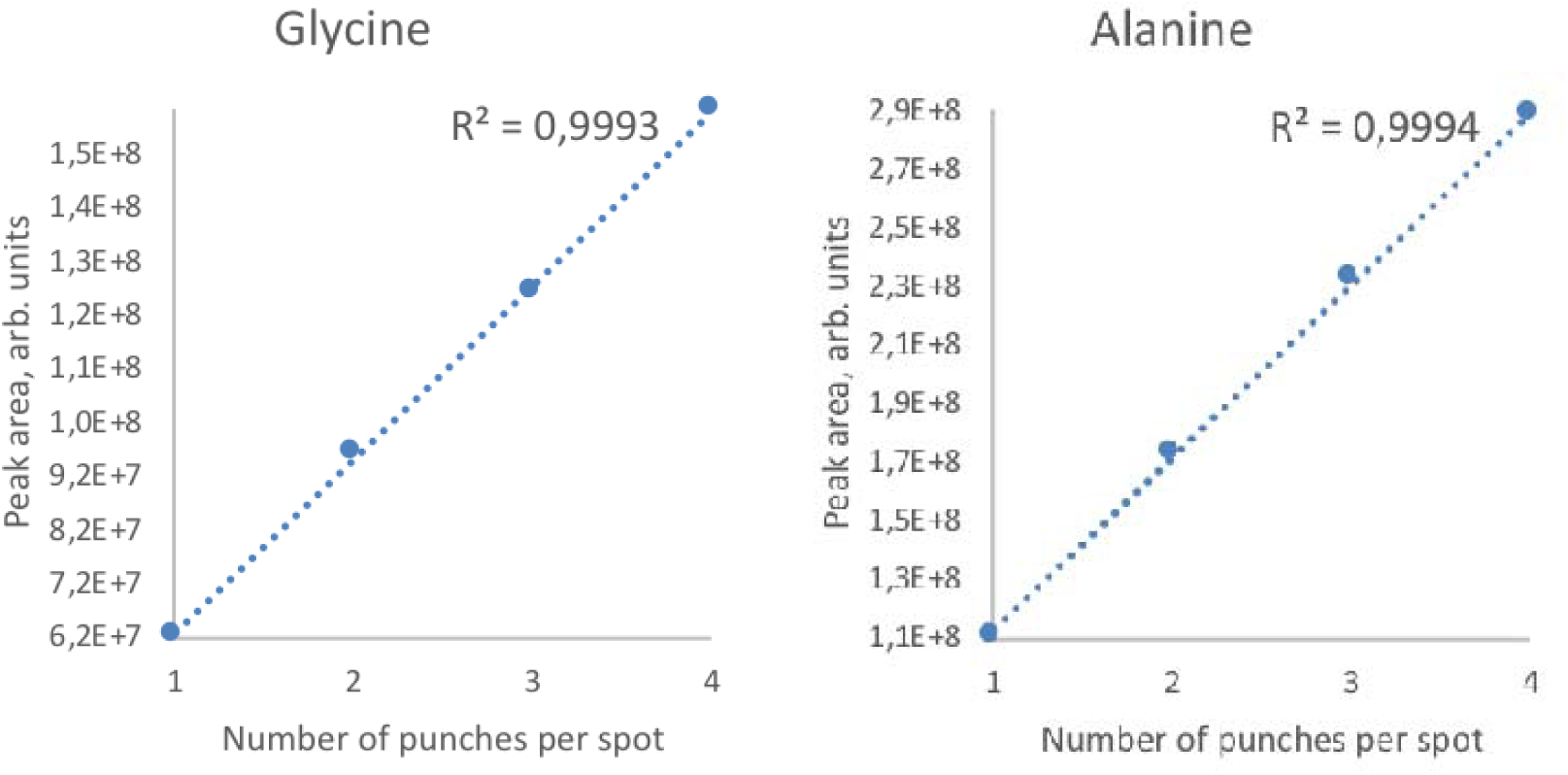
Linear increase of peak area with increasing number of punches from one spot for glycine and alanine.

### 3.3 Application/proof of concept

#### 3.3.1 Targeted approach

The method covers a large part of the metabolome. Endogenous metabolites ranging in polarity from logP −4.4 to 8.8 are readily detected; see **Table 2** for a list of representative compounds, highlighted here as they are all employed as biomarkers in e.g. newborn screening, and for maple syrup urine disease and tyrosinemia (among other diseases). In addition, **Figure 8** shows a total ion chromatogram from a positive ionization dried blood spot analysis with mass spectra and extracted ion chromatograms of eight selected detected endogenous compounds, illustrating the even peak distribution of hydrophilic and hydrophobic compounds along the chromatogram.

**Table 2.** Examples of detected endogenous compounds with a wide range in polarity. LogP values were obtained from Pubchem [27].

| Amino acid | Chemical formula | Exact mass, Da | LogP | Acylcarnitine | Chemical formula | Exact mass, Da | LogP |
| --- | --- | --- | --- | --- | --- | --- | --- |
| **Ornithine** | $C_5H_{12}N_2O_2$ | 132.090 | -4.4 | **C0** | $C_7H_{15}NO_3$ | 161.106 | -0.2 |
| **Citrulline** | $C_6H_{13}N_3O_3$ | 175.096 | -4.3 | **C2** | $C_9H_{17}NO_4$ | 203.116 | 0.4 |
| **Arginine** | $C_6H_{14}N_4O_2$ | 174.112 | -4.2 | **C3** | $C_{10}H_{19}NO_4$ | 217.131 | 0.9 |
| **Glycine** | $C_2H_5NO_2$ | 75.032 | -3.2 | **C4** | $C_{11}H_{21}NO_4$ | 231.147 | 1.2 |
| **Alanine** | $C_3H_7NO_2$ | 89.048 | -3.0 | **C5** | $C_{12}H_{23}NO_4$ | 245.163 | 1.8 |
| **Valine** | $C_5H_{11}NO_2$ | 117.079 | -2.3 | **C8** | $C_{15}H_{29}NO_4$ | 287.210 | 3.4 |
| **Tyrosine** | $C_9H_{11}NO_3$ | 181.074 | -2.3 | **C12** | $C_{19}H_{37}NO_4$ | 343.272 | 5.5 |
| **Methionine** | $C_5H_{11}NO_2S$ | 149.051 | -1.9 | **C14** | $C_{21}H_{41}NO_4$ | 371.304 | 6.6 |
| **Leucine** | $C_6H_{13}NO_2$ | 131.095 | -1.5 | **C16** | $C_{23}H_{45}NO_4$ | 399.335 | 7.7 |
| **Phenylalanine** | $C_9H_{11}NO_2$ | 165.079 | -1.5 | **C18** | $C_{25}H_{49}NO_4$ | 427.366 | 8.8 |

**Figure 8.**
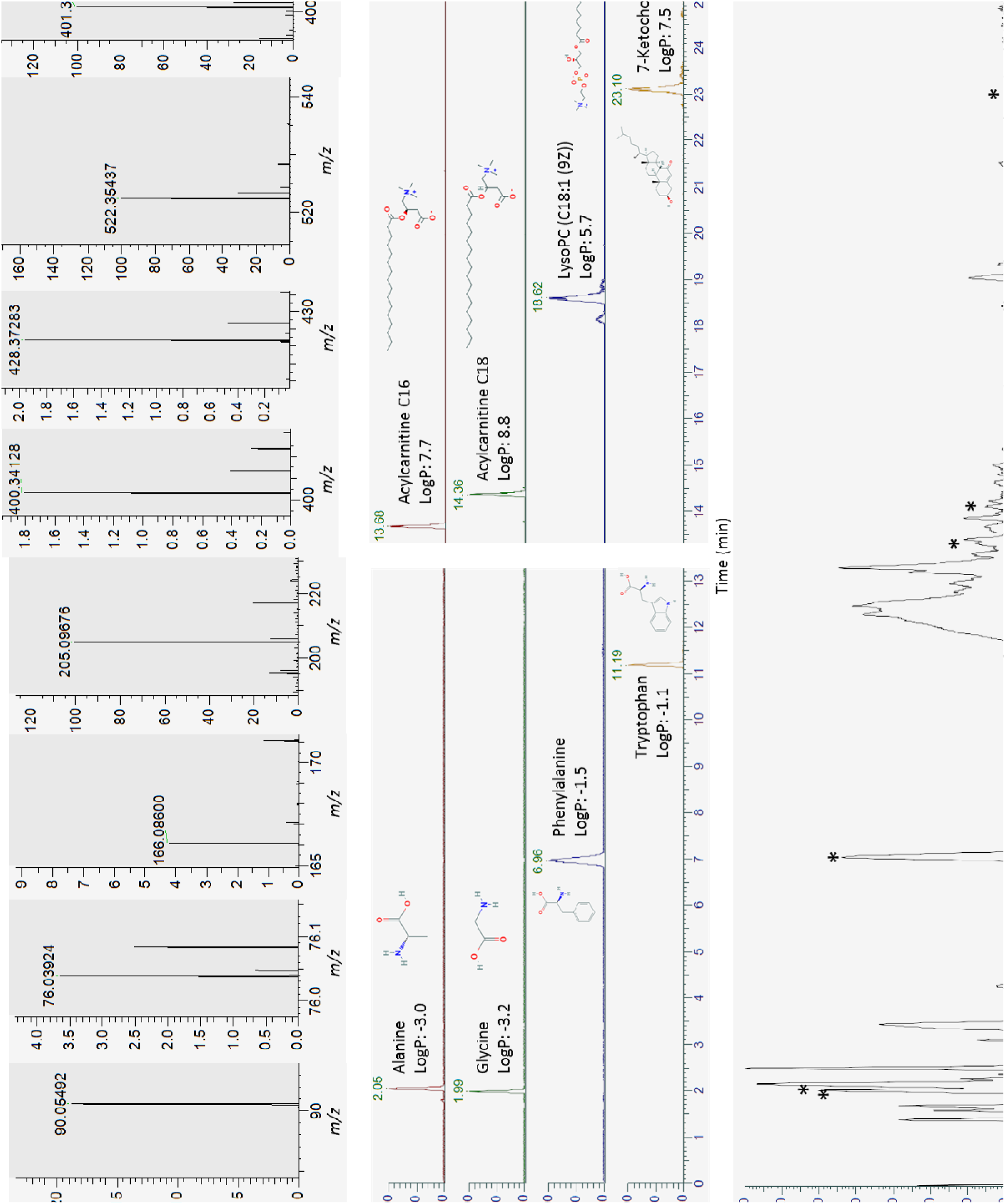
Mass spectra (A) and extracted ion chromatograms with names, logP values and structures (B) of a selection of detected endogenous compounds in a dried blood spot (alanine, glycine, phenylalanine, tryptophan, acylcarnitine C16, acylcarnitine C18, LysoPC (C18:1 (9Z)) and 7-ketocholesterol), and total ion chromatogram (C) with asterisks indicating at which points in the chromatogram the spectra in A are located. LogP values and structures were obtained from Pubchem [27].

We also investigated if our method could detect changes in the concentration of one target metabolite out of the thousands of metabolites detected. Six healthy volunteers were taken DBS samples of during free intake of coffee and soda, and during no intake of coffee and soda. Caffeine is a suitable compound to monitor as we know that it is exogenous (mostly originating from coffee and soda), and people consume various amounts of the substance. Thus, caffeine was measured in all samples. As shown in **Figure 9**, the measured amount of caffeine decreased with increasing time since intake.

**Figure 9.**
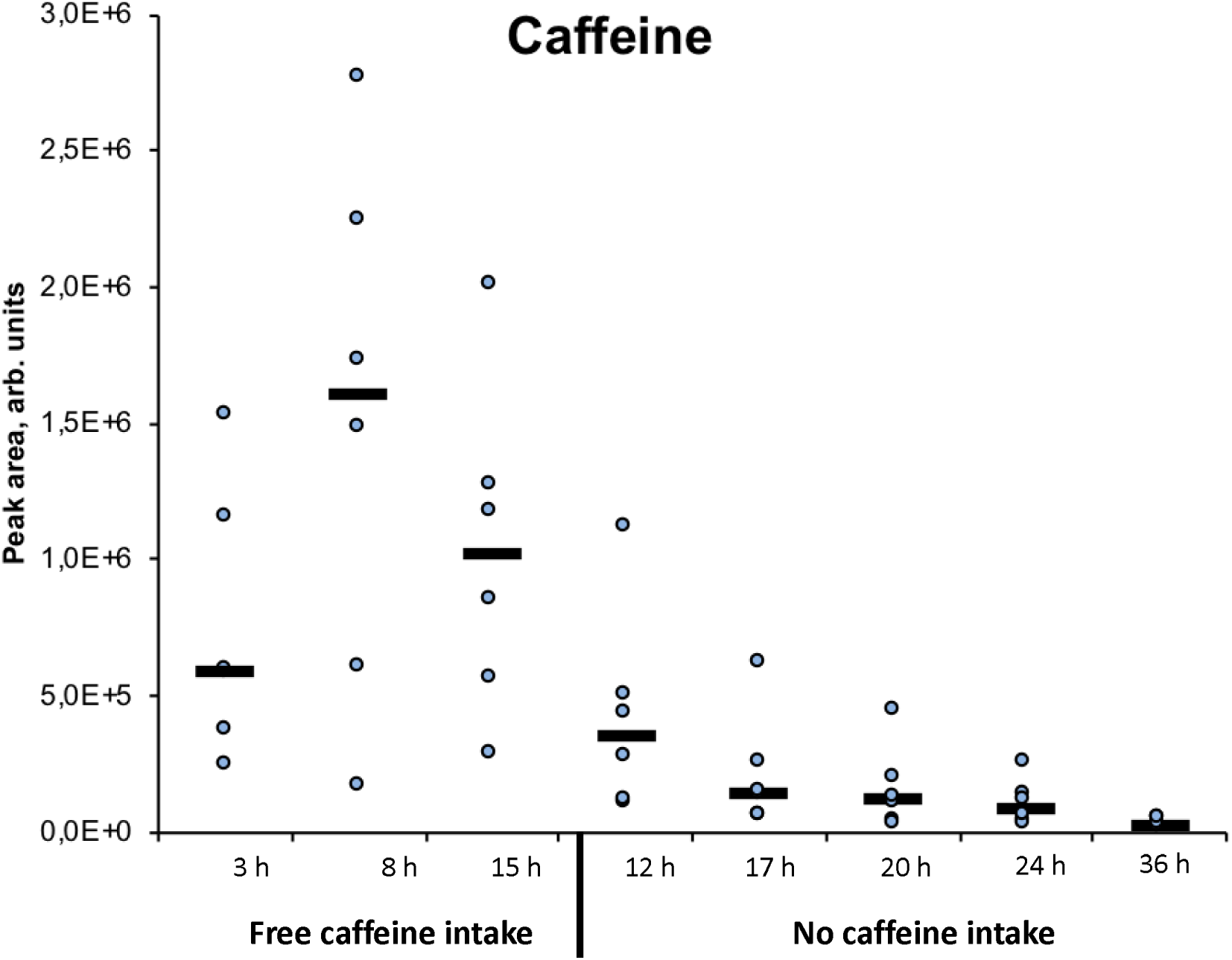
Measured amounts of caffeine decreased with time since intake. The spread in measured peak area is likely caused by different intakes of coffee between the participants.

#### 3.3.2 Detection of differences in nutritional states (untargeted analysis)

An additional proof of concept study was performed by analyzing DBS samples from six healthy volunteers during free food intake and during fasting. DBS samples were taken during free diet, after 12 hours of fasting and after 36 hours of fasting. The volunteers were allowed to drink as much water as they wanted during the fasting period. A principal component analysis plot of samples taken from all volunteers during free diet and after 12 hours and 36 hours of fasting, respectively, is shown in **Figure 10**. The samples from the three nutritional states clearly grouped as three separate clusters, with the apparent exception of the free diet sample from person F, which clustered together with samples taken after 12 hours overnight fasting. However, it turned out that person F was actually omitting breakfast that day, meaning that this point is correctly located together with the overnight fasting samples and should in fact be classified as an overnight fasting sample.

**Figure 10.**
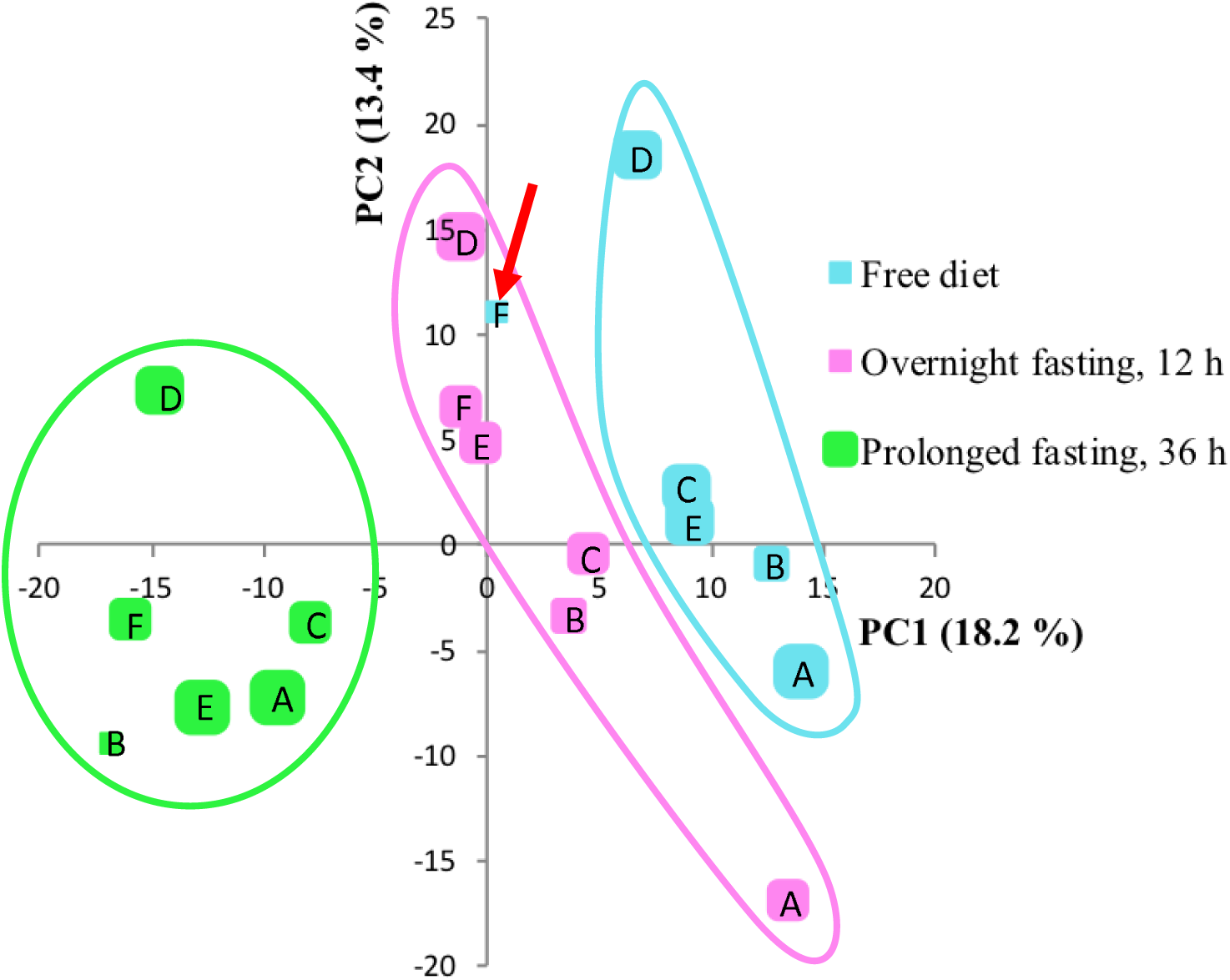
Principal component analysis plot of DBS samples from six individuals (A-F) taken after free diet, overnight fasting (12 hours) and prolonged fasting (36 hours). One point represents metabolites detected in that particular sample. Individual F did not eat before collecting the free diet sample. This point (red arrow) thus clustered correctly together with the overnight fasting samples.

To evaluate the method’s ability to identify discriminating compounds between groups, we used a volcano plot to compare samples taken after overnight fasting (12 hours) with samples taken after prolonged fasting (36 hours) (**Figure 11**). Compounds with a significantly lower concentration in prolonged fasting samples were identified: caffeine, theobromine, and paraxanthine, all associated with metabolism of (coffee) drinks [28,29]. An upregulated marker of prolonged fasting samples was identified as β-hydroxybutyrate, a ketone body naturally produced during fasting for energy transfer [30]. Taken together, the single run platform was well suited for revealing trends of expected metabolism in this controlled study.

**Figure 11.**
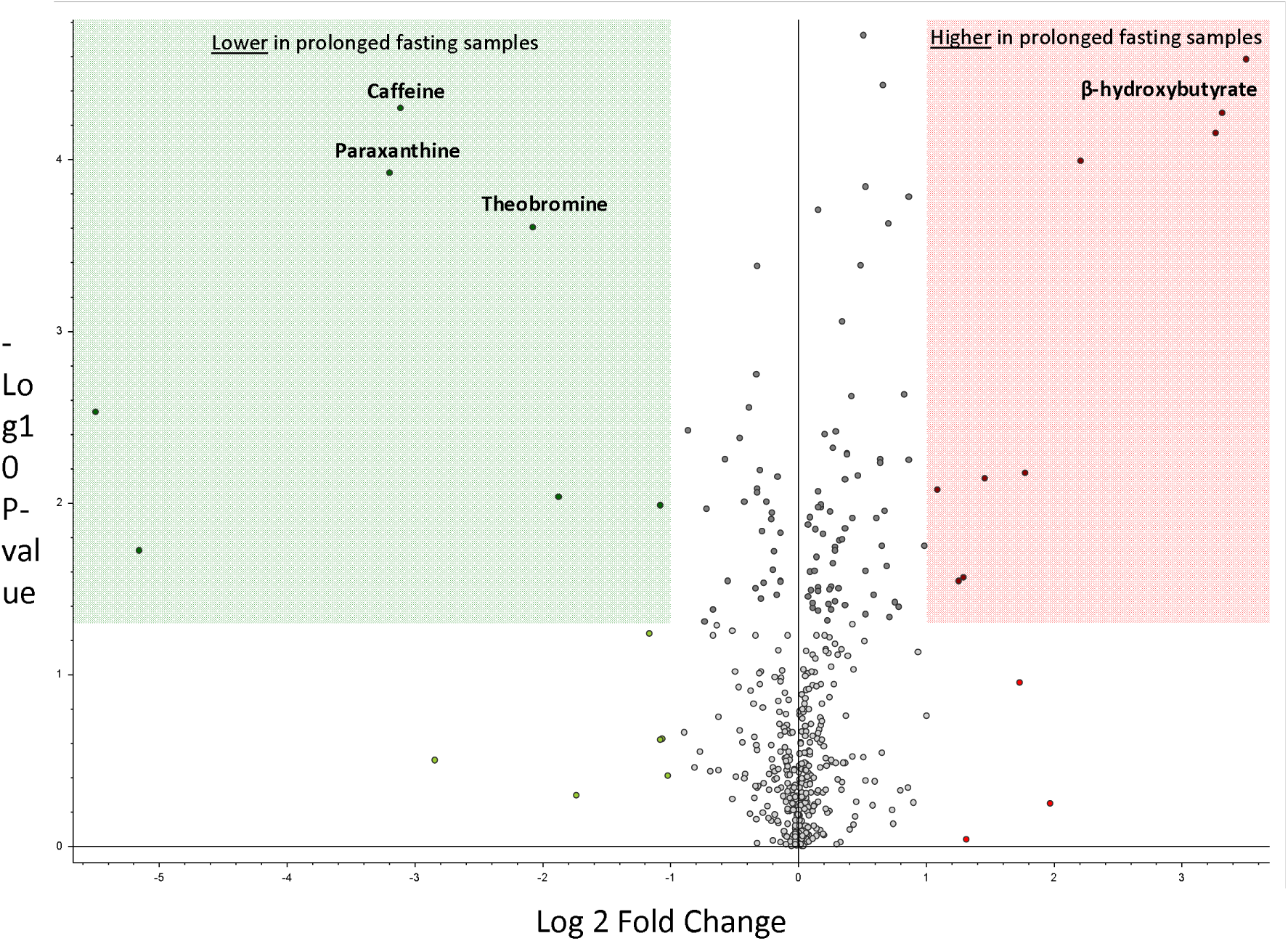
Volcano plot of DBS samples from six individuals taken after overnight fasting (12 hours) and prolonged fasting (36 hours). One point represents one compound. Green box: lower concentration measured in samples taken after 36 hours than after 12 hours of fasting. Red box: higher concentration measured in samples taken after 36 hours than after 12 hours of fasting.

## 4 Conclusions

A single LC-MS method has satisfactory analytical performance for a broad range of metabolites with regards to polarity, having evaluated and optimized parameters regarding MS sensitivity, column separation and sample preparation, demonstrated with proof-of-concept studies regarding both untargeted and targeted metabolite approaches using the same method/settings. We find that a single LC-MS method can indeed be a compromise between multi-method deep profiling and fast “shotgun” approaches. We have here evaluated our platform with regards to key biomarkers of inborn errors of metabolism, and are currently exploring the limitations of our platform regarding lipids such as phosphatidylcholines, cholesterol esters, and acylglycerols (employing SPLASH^®^ standards).

## Data Availability

The data presented and described in this manuscript is from liquid chromatography-mass spectrometry recordings. The raw data files are available upon request.

## Acknowledgements

Financial support from UiO:Life Science is gratefully acknowledged. This work was also partially supported by the Research Council of Norway through its Centre of Excellence scheme, project number 262613. S.R.W. is a member of the National Network of Advanced Proteomics Infrastructure (NAPI), which is funded by the Research Council of Norway INFRASTRUKTUR-program (project number: 295910).

