## Supplemental Material for "Bridging the polar and hydrophobic metabolome in single-run untargeted liquid chromatography-mass spectrometry dried blood spot metabolomics for clinical purposes"

**Table 1.** Average peak area (PA) and retention time (RT) relative standard deviation (RSD %) of selected compounds measured in a DBS injected three times each day during an analysis run of 11 days.

| **Compound** | **Average PA (a.u.) N=30** | **RSD % PA (a.u.) N=30** | **Average RT (min) N=30** | **RSD % RT (min) N=30** |
| --- | --- | --- | --- | --- |
| **Ornithine** | 1.81E+07 | 4 % | 1.62 | 0.3 % |
| **Arginine** | 6.88E+06 | 3 % | 1.75 | 0.4 % |
| **Citrulline** | 1.75E+07 | 3 % | 2.19 | 0.2 % |
| **Valine** | 1.71E+08 | 9 % | 2.50 | 0.1 % |
| **Methionine** | 8.17E+06 | 7 % | 3.05 | 0.2 % |
| **Leucine** | 1.37E+08 | 6 % | 3.33 | 0.3 % |
| **Tyrosine** | 3.34E+07 | 5 % | 4.10 | 0.3 % |
| **Phenylalanine** | 6.73E+07 | 3 % | 6.73 | 0.2 % |
| **Acylcarnitine C0** | 5.16E+07 | 2 % | 2.11 | 0.3 % |
| **Acylcarnitine C2** | 3.10E+07 | 4 % | 2.61 | 0.2 % |
| **Acylcarnitine C3** | 1.02E+06 | 8 % | 4.97 | 0.4 % |
| **Acylcarnitine C8** | 2.09E+05 | 8 % | 12.09 | 0.1 % |
| **Acylcarnitine C14** | 3.35E+05 | 10 % | 13.14 | 0.1 % |
| **Acylcarnitine C16** | 2.90E+06 | 4 % | 13.69 | 0.1 % |
| **Acylcarnitine C18** | 1.14E+06 | 4 % | 14.42 | 0.1 % |
